# A simple tool for Visualizing Time Sections of Transesophageal Echocardiography with Python

**DOI:** 10.1101/2024.11.26.24317630

**Authors:** Marc Fiammante, Pierre Dellamonica, Emilie Mertens, Arnaud De La Chapelle, Laury Leveille, Mohamed Labbaoui

**Affiliations:** Unaffiliated, Independent Researcher, Retired IBM Fellow; Infectious Disease Specialist, Université Côte d’azur; Cardiologist ultrasound specialist, Arnault Tzanck Institute Saint Laurent du Var; Anesthesiologist resuscitator, Arnault Tzanck Institute Saint Laurent du Var; Adult Cardiac Surgeon, Arnault Tzanck Institute Saint Laurent du Var

**Keywords:** Transesophageal echocardiography (TEE), Infectious Endocarditis, Ultrasound video, Cardiology, Vegetations, Optical Flow, Plotly, Dash

## Abstract

**Background:** Transesophageal echocardiography (TEE) is a critical tool in diagnosing and managing infectious endocarditis, providing detailed images of cardiac structures. However, identifying vegetations on valves and their dynamic behavior in ultrasound videos can be challenging. TEEs metadata often does not include scale enabling computation of speed.

**Objectives:** To address this, we developed a simple Python-based tool that enhances the visualization of these dynamic characteristics. This tool reconstructs an optical flow from TEE images, capturing the motion of cardiac structures and offering deeper insights into their behavior. The tool also recovers scale from visual information on the TEES.

**Methods:** By leveraging the Marching Cubes algorithm and 2D Fast Fourier Transform (FFT) to recover scale from images, the tool efficiently processes video frames to create a 3D representation where time is the third dimension. Wit his mouse the user can select temporal slices and a view of the dynamic evolution in that slice is created together with the speeds.

**Results:** This approach allows for measurement of thicknesses and speeds, aiding in the evaluation of valvular and vegetation dynamics.

**Conclusions:** The tool’s user-friendly interface, built with Dash and Plotly, enables interactive analysis and visualization, making it a valuable asset for cardiologists in clinical settings to further analyze valvular behavior.

## Introduction

Transesophageal echocardiography (TEE) is a critical imaging technique used by cardiologists to obtain detailed images of the heart and its structures. Traditional TEE provides two-dimensional images, but there is a growing need for tools that can visualize the temporal evolution of these images, effectively creating a three-dimensional view where the third dimension is time. This article introduces a simple Python-based tool designed to help cardiologists visualize time sections of TEE, applicable to a variety of ultrasound device brand, enhancing their ability to diagnose and monitor valvular cardiac conditions.

### Clinical need and use

Transesophageal echocardiography (TEE) is a specialized form of echocardiography that provides detailed images of the heart and its structures by using an ultrasound transducer positioned in the esophagus. This proximity allows for clearer and more precise images compared to transthoracic echocardiography (TTE), especially for certain cardiac conditions.

The development of TEE began in the 1970s with the use of esophageal ultrasound to measure blood flow in the aortic arch. By the 1980s, flexible probes with phased-array transducers were introduced, allowing for more detailed imaging. . The evolution continued with the development of biplane and multiplane probes, and more recently, real-time three-dimensional (3D) imaging capabilities [1 ].

Transesophageal echocardiography is an indispensable tool in the diagnosis and management of infectious endocarditis. Its ability to provide detailed and accurate images of cardiac structures makes it superior to TTE in many clinical scenarios. By detecting vegetations, assessing prosthetic valves, identifying complications, and guiding surgical interventions, TEE significantly contributes to the effective management of this serious condition [2].

When vegetations on heart valves are detected using Transoesophageal Echocardiography (TEE), the decision to proceed with surgery involves several critical steps and considerations, including a good evaluation of vegetations size, shape and dynamic behaviour. Large vegetations with a small attachment to the valve are associated with a higher risk of embolic events and complications [3].

There are relatively few articles that specifically focus on the detailed evaluation of the shape, speed, and size of vegetations attached to cardiac valves. Most research tends to emphasize the size of vegetations, some mention oscillations and their clinical implications, such as the risk of embolic events and the need for surgical intervention [4] [5] [6] [7].

Some research have created computational models based on TEEs to recover kinematic and anatomical information in the form of left heart chambers and valve boundaries through a level-set-based, user-in-the-loop segmentation on 2-D and 3-D TEE (8]. User-in-the-loop for segmentation of 3D TEEs is a rather lengthy process and there is a need for a simpler and lighter approach.

### Purpose and overview of the Tool

There are existing tools for cardiac image analysis and modeling [14]. The purpose of the tool is to provide a simple mean for enabling a better evaluation of valvular and vegetation size and dynamic behavior using Transesophageal echocardiography (TEE) resulting video clips. This tool reconstructs an optical flow from TEE images that can enhance the analysis by capturing the motion of cardiac structures, offering deeper insights into dynamic behavior. Optical flow refers to the pattern of apparent motion of objects, surfaces, and edges in a visual scene, caused by the relative motion between an observer and the scene. In the context of echocardiography, optical flow can be used to track the movement of cardiac tissues, hopefully providing valuable information about heart dynamics. A specific requirement was the ability to analyze thicknesses and relative speeds between valves and vegetations.

### Selecting an optical flow algorithm usable with images from TEE videos

Advanced optical flow algorithms, such as the Lucas-Kanade or Horn-Schunck methods, involve complex mathematical computations that require substantial processing power and after testing we made out that they are not usable in the context of a fast analysis. Some authors even propose hardware implementation to alleviate the need for processing power [9].

In consequence our approach has been to define a faster approach that can reconstruct an optical flow at low computing processing power and allow an easy visual manipulation of the result. An optical flow is a mean of creating a 3D flow from a set of video frames where the third dimension is the time. There is an algorithm in the spatial domain called the Marching Cubes algorithm which is a method for creating a 3D shape from a stack of 2D slices (like layers in a CT scan or MRI) [15]. Imagine you have a grid of cubes inside an object. The algorithm “marches” through each cube to check if the object’s surface passes through it. Based on where it intersects, each cube is given a pattern that represents parts of the surface. These patterns are used to draw small triangles that, when combined, form a smooth 3D model of the entire object’s surface. This method is often used in medical imaging and 3D graphics because it helps build detailed and accurate 3D shapes from simpler data.

We just replaced the spatial slices by temporal slices which are the echography video frames and it resulted in a fast computation of an optical flow.

The mesh vertices and faces result from the marching_cubes method of the skimage.measure module where the optimal luminosity level we tested is 92 in a range of 0 to 255. We selected Plotly [12] as the most practical mean to display the results.

The figure 1 shows on the right the mesh displayed from the TEE on the left with Plotly graphic object Mesh3D (a rainbow coloring is used to differentiate frames). In the application the 3D view can be zoomed and rotated with the mouse.

**Fig 1.**
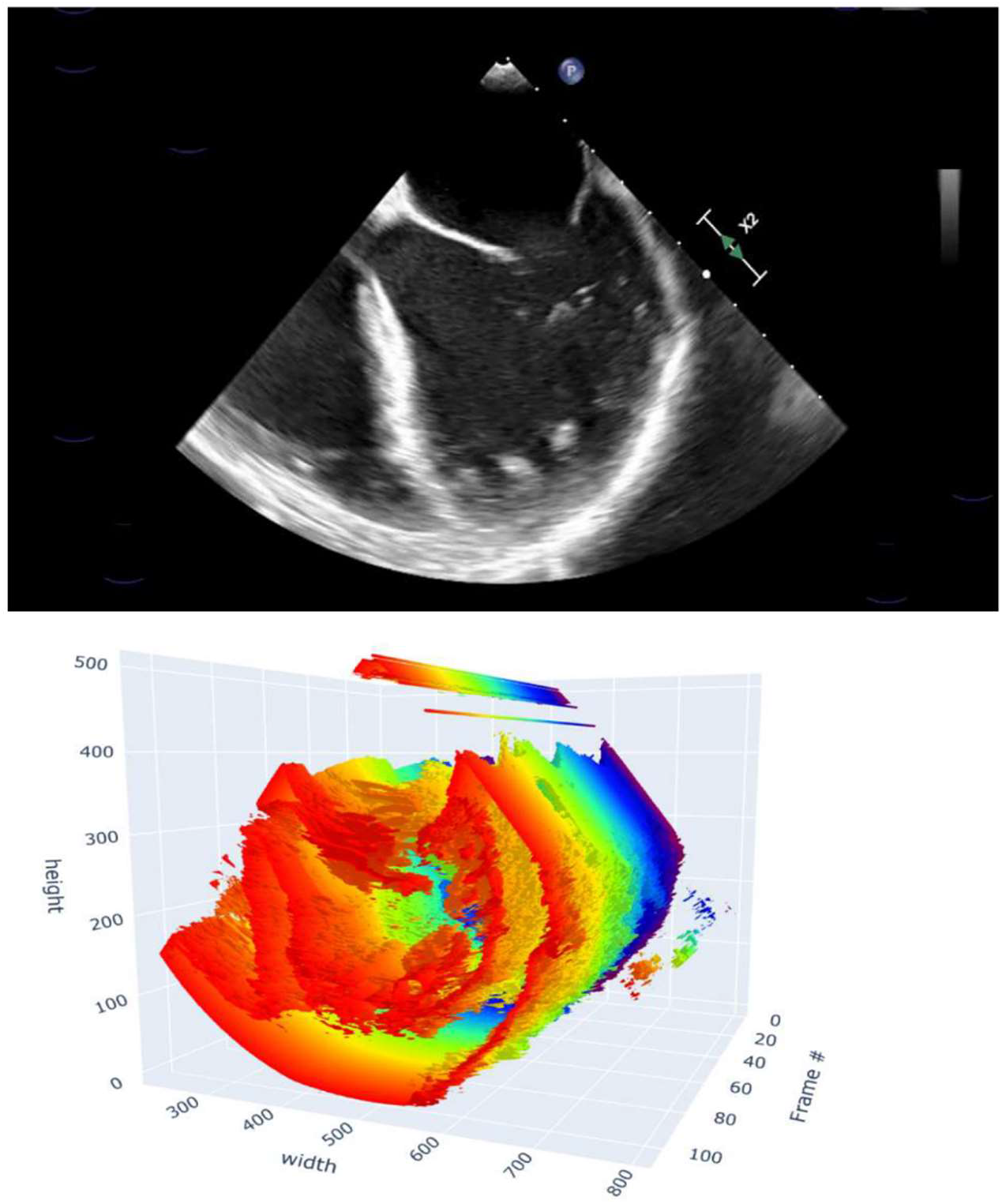

### Recovering time and scale

For purpose of measuring thicknesses and speed it is essential to recover the elapsed time between two frames and the scale in pixels per centimeter.

Even though Dicom and Nifti format metadata can carry resolution or duration, but this information was not present in the files exported from our systems and we had to revert to another approach by using the information directly from the video files. We could not locate any TEE files in the open source, but to confirm we checked that scale was present but not duration information was not available from Nifti files of the Camus database [10], and similarly Echonet Dynamic dataset [11] contains AVI video files and informational csv which do not have a pixel per cm scale information available.

So our approach was to revert to information available in the video files, the frames per second for the time information and the scales visible on the images either composed of white dots or colored dashes on the images exported from the various ultrasound device brands. Elapsed time is rather easy as mp4 or AVI videos carry in their metadata the frames per second information. Recovering scale however happened to be more a difficult problem as the information from the transducers is lost in the export.

The approach that worked in our case is the 2D Fast Fourier Transform (FFT) which is a powerful tool used in image processing to analyze the frequency content of an image. Here’s a brief overview of how it works and its applications [16][17]:

The 2D FFT transforms an image from the spatial domain (where pixel values represent intensity) to the frequency domain (where pixel values represent frequency components). This transformation helps in understanding the periodic structures within the image. Frequency Domain Representation: The resulting 2D array contains complex numbers representing the amplitude and phase of the frequency components. In the frequency the low frequencies: represent the regularly distant spaced, smooth or gradual changes in the image (e.g., background or scale dots) while the high frequencies represent the sharp, abrupt changes (e.g., edges, noise). Filtering out the center components of the frequency domain by zeroing a square zone of plus and minus 100 of the frequency domain only leaves the dots or dashes. Then a polar warp centered on the transducer location and a peaks analysis results in the pixel/cm scale as the dots or dashes are spaced every centimeter.

The figure 2 shows four images from the scale recovery process sequence using the 2D Fast Fourier Transform to isolate scale.

**Fig 2.**
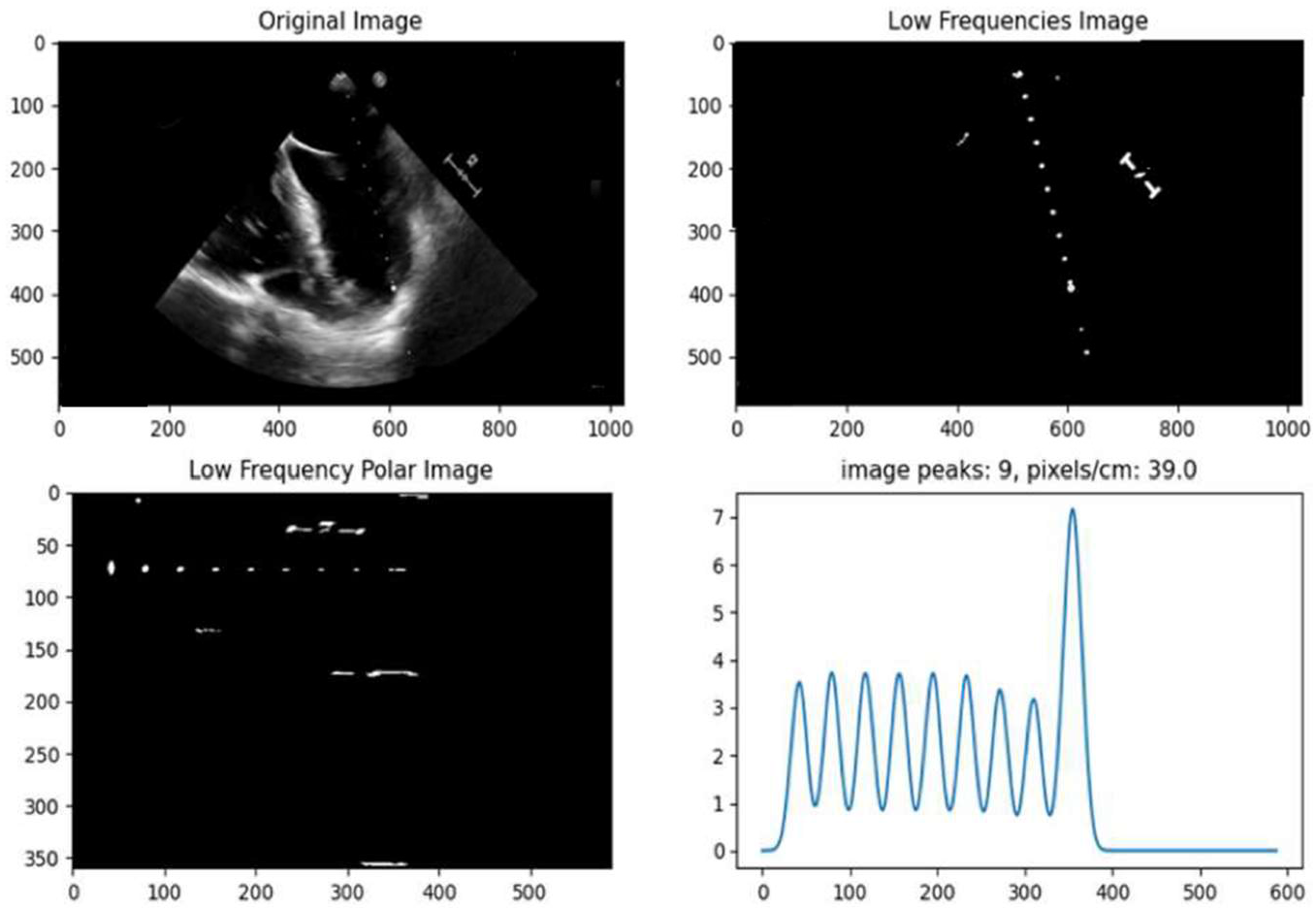

### Selecting the slice for temporal analysis

We use Dash to create a local applications for viewing and analyzing data on the cardiologist standalone computer, without any network exposure. When configured this way, the application runs locally, using the computer’s resources to process and display data. The layout, defined in Python, allowed us to set up interactive elements to select a slice of the TEE either with a rectangle or line selection. When rectangle selection occurs, a callback computes masks the frames to leave only the zone of the rectangle for 3D visualization and the longest midline segment for computing the intersection of the 3D mesh with a plane with same normal as the segment to create a 2D slice across all frames containing the segment, the vertical abscissa being the segment and horizontal the time.

When line selection occurs resulting in a segment, a mask is created with the segment and a few pixels around to create the 3D visualization respecting the angle of the line, and the intersection across the 3D mesh is computed using a plane which normal is the normal of the segment.

The figure 3 shows the 3D image with time as depth resulting from rectangular selection around a particular zone such as a valve on the TEE

**Fig 3.**
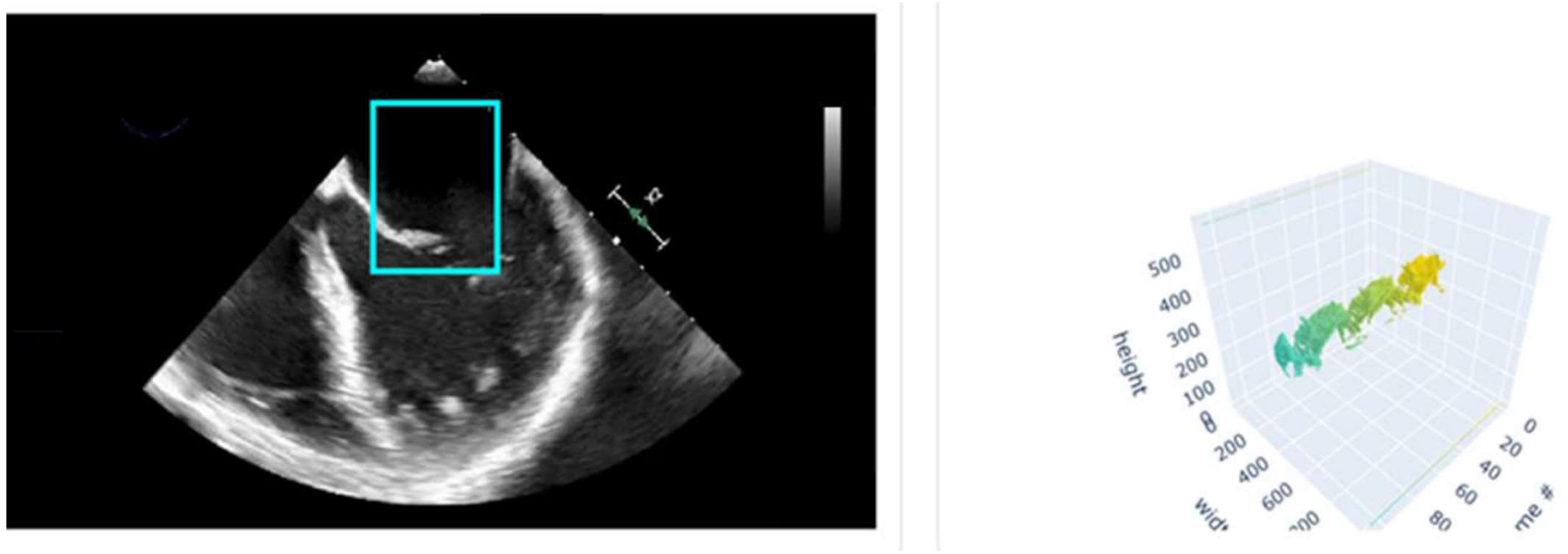

The figure 4 illustrates the line or rectangle selection options, with the line permitting sections with any angle

**Fig 4.**
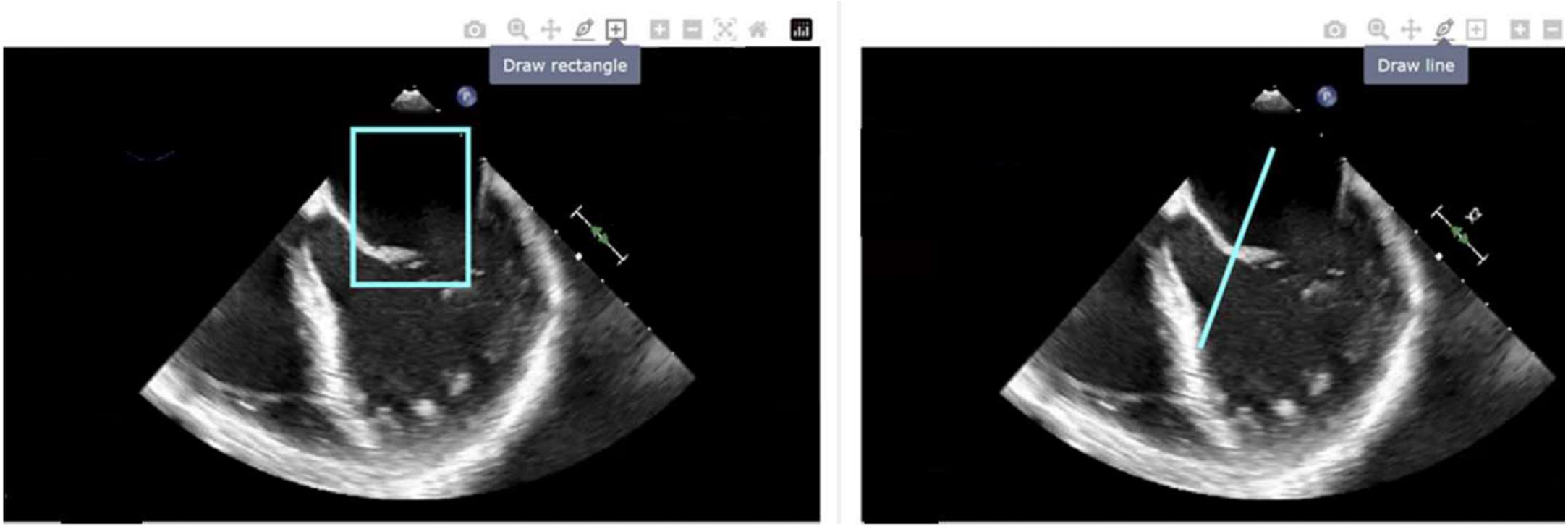

The polygons from the 2D slice are retrieved, and the gradients computed to result in the speed in centimeter per second at each point of the polygons. Three views are constructed on a page division below the previously discussed TEE and 3D views with time and speed information. The left pane displays the time section of the selected zone, and Dash application visualization capabilities provides Plotly flyovers that display the points in centimeter, together with time coordinates and computed speed. The middle pane retains major boundaries movements, smoothed for an average speed computation and the right pane displays the corresponding speed curves with identical trace numbering.

In a second pane polygons are split by sections that are not wrapped and smoothed to simplify the view with identifiable traces in a legend. The third panel is a speed curve view when to the user views the speed curve for each trace.

The figure 5 shows the application page row with the three panes resulting from the selection. The left pane with the time evolution of the boundaries, here vegetation and mitral valve boundaries, the second pane has the polygons a split by smoothed and growing time sections with identifiable traces in a legend, traces can be individually isolated and shown. The third panel is a speed curve where the user can also isolate and show specific traces, which have the same number as in the second pane.

**Fig 5.**
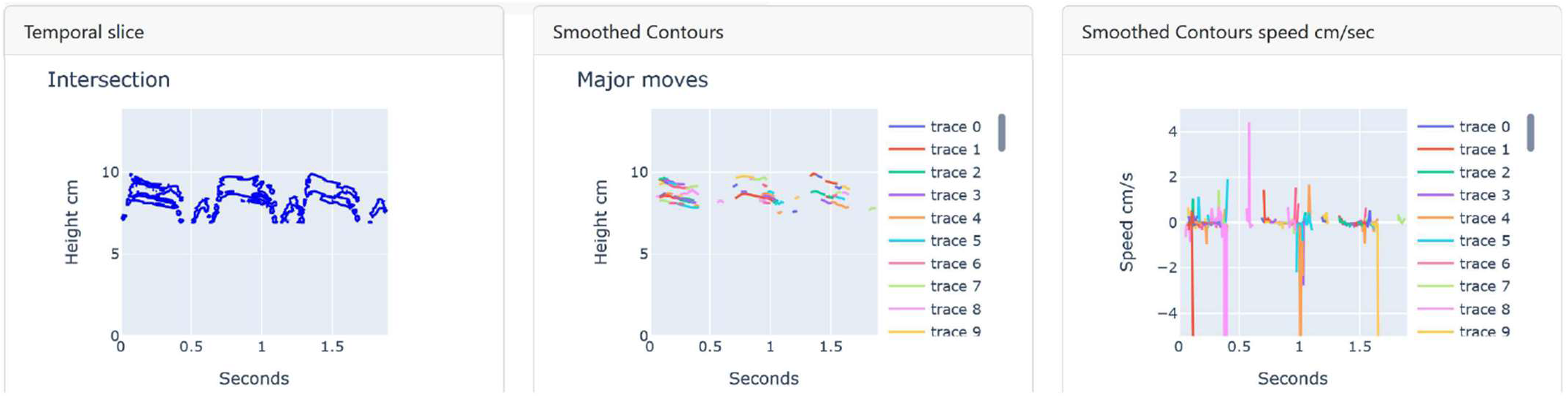

On the smoothed view given traces can be selected by double clicking on its legend label as well as on the speed curve view.

The figure 6 is an example result of the trace selection to isolate specific boundaries to analyze respecting speeds.

**Fig 6.**
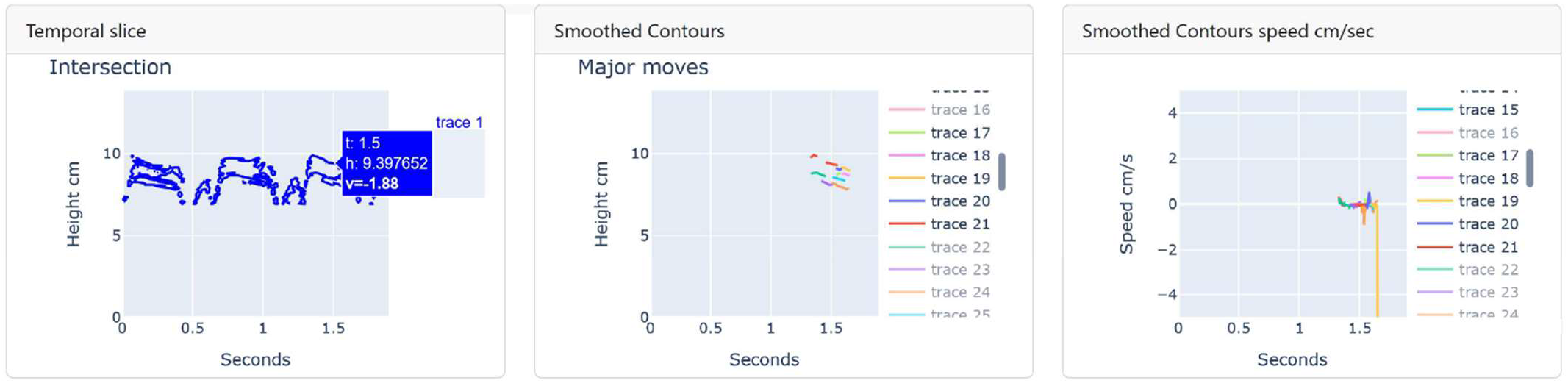

### Technical Implementation

The user interface is displayer in a Web browser interacting with a local lightweight web server started by Dash. The server runs on the localhost IP address, not opening any external port thus limiting the interactions to the machine without any cyber risks.

The overall application layout is shown on the figure 7. Targeting simplicity only one page contains all of the necessary information.

**Fig 7.**
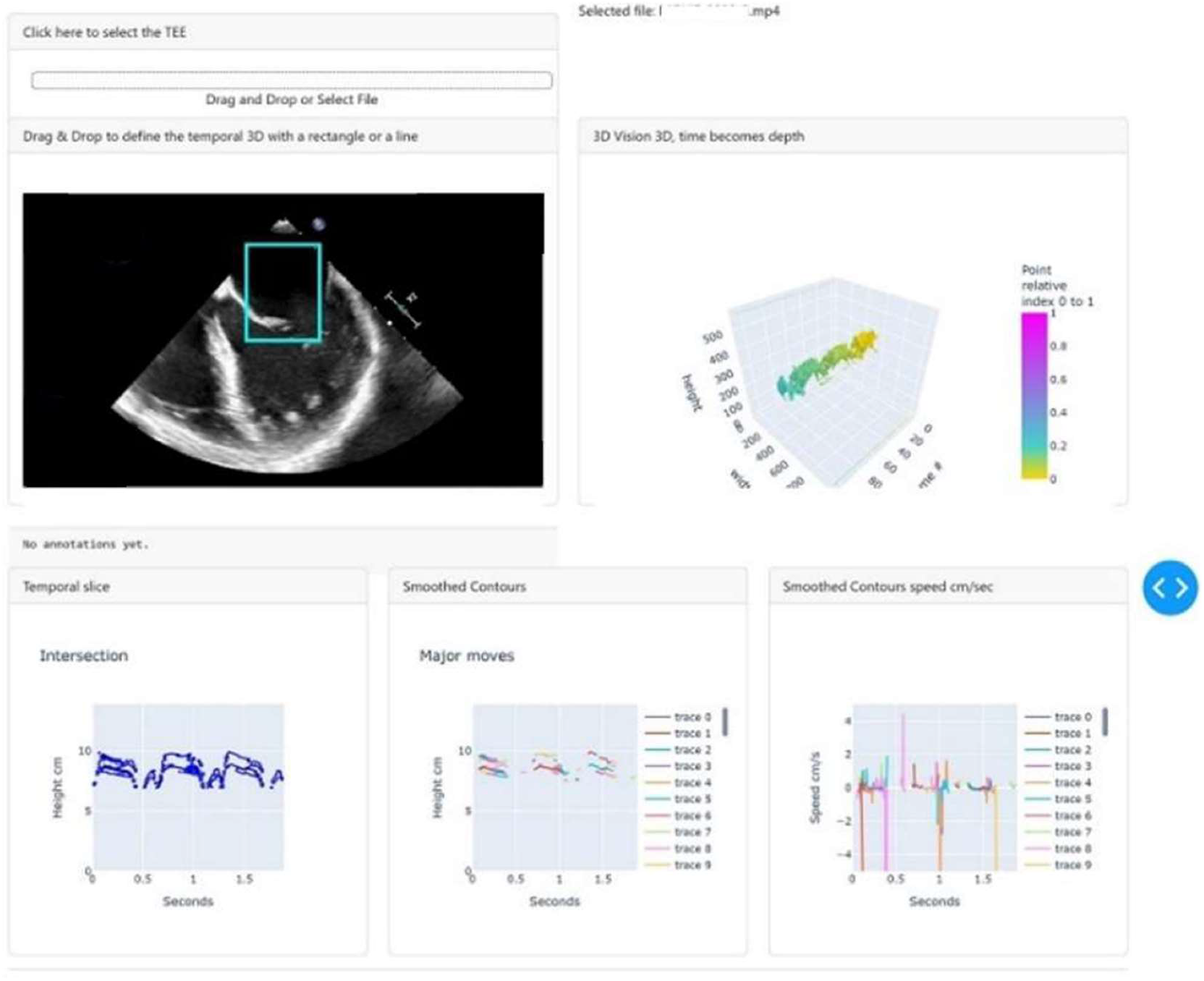

The code leverages several Python libraries to achieve its functionality:

1. **Imageio [18], skimage [19], PIL [20], pymediainfo [21], trimesh [22], and scipy [23]**, : For video and image processing tasks, including fft, frame manipulation, masking, polygon approximation, marching cubes, 3D mesh operations and scale recovery.
2. **Dash [24], and Plotly [25]:** For the application layout and interaction, drag and select out of the box, creating the visualizations and plotting the time evolution of the selected sections.

These libraries enable a low code implementation with less than 800 lines of Python.

## Conclusion

This Python tool represents a significant advancement in the visualization of TEE data. By providing a 3D view where time is the third dimension, it allows cardiologists to gain a deeper understanding of cardiac valve dynamic behavior. The ability to recover scale accurately and interactively select areas of interest makes this tool both powerful and user-friendly. As cardiac imaging continues to evolve, tools like this should play a role in improving diagnosis and patient outcomes.

## Data Availability

All data produced in the present study are available upon reasonable request to the authors

## Acknowlegements

We thank Olivier Casile for his help with the overall problem analysis.

## Notes

### Competing Interest Statement

The authors have declared no competing interest.

### Funding Statement

This study did not receive any funding

### Author Declarations

Ethics committee/IRB of Institut Arnault Tzanck, St Laurent du VAR, France gave ethical approval for this work . Signed by Michel Salvadori General Director of Institut Arnault Tzanck.

